# Estimated prevalence of multiple chronic conditions throughout adulthood using data from the *All of Us* Research Program

**DOI:** 10.1101/2024.10.17.24315661

**Authors:** Xintong Li, Caitlin Dreisbach, Carolina M. Gustafson, Komal Patel Murali, Theresa A. Koleck

## Abstract

Estimation of multiple chronic condition (MCC) prevalence throughout adulthood provides a critical reflection of MCC burden. We analyzed electronic health record codes for 58 conditions to estimate MCC prevalence for *All of Us* (*AoU*) Research Program adult participants (N=242,828). Approximately 76% of *AoU* participants were diagnosed with MCCs, with over 40% having 6 or more conditions and prevalence increasing with age; the most frequently occurring MCC combinations varied by age category (i.e., mental health conditions in early adulthood and physical health conditions in middle adulthood through advanced old age). We report notable prevalence of MCC throughout adulthood and variability in MCC condition combinations by age category in *AoU* participants. These findings highlight the need for targeted, innovative care modalities and population health initiatives to address MCC burden throughout adulthood.

## Introduction

Multiple chronic conditions (MCCs) is defined as the presence of two or more chronic conditions such as Alzheimer’s disease, arthritis, asthma, cancer, chronic kidney disease, diabetes, heart failure, and hypertension (1). Approximately 80% of adults 65 years of age and older in the United States (U.S.) are living with MCCs (2). Diagnosis with MCCs, however, is not limited to older adults. One study estimated that 22% of young adults, 18-34 years of age, are diagnosed with MCCs (3). Moreover, research suggests that more contemporary generations of adults have greater MCC burden and are diagnosed with MCCs at earlier ages than previous generations (4). Estimation of the prevalence of MCCs throughout all stages of adulthood is a critical reflection of MCC burden; it is important to examine the prevalence of MCCs broadly using regularly updated data sources to inform targeted prevention and management strategies as well as resource prioritization. Thus, the purpose of this study is to estimate the prevalence of MCCs for adult participants in the National Institutes of Health *All of Us* (*AoU*) Research Program dataset – a curated disease and population-agnostic, longitudinal biomedical dataset of diverse individuals living in the U.S. – to gain a deeper understanding of MCCs throughout adulthood (i.e., early adulthood, middle adulthood, late-middle adulthood, late adulthood, and advanced old age).

## Materials and Methods

We conducted a cross-sectional analysis using the *AoU* Controlled Tier version 7 data release (date of last access 16/06/2024) in the secure, cloud-based *AoU* Researcher Workbench platform. This study was exempt from human subjects’ approval as only deidentified data were analyzed. Using the Cohort Builder tool, we identified all adult participants 18 years of age or older who consented to share electronic health record (EHR) data and had at least one International Classification of Diseases-10th Revision (ICD-10) code documented to ensure the availability of EHR data (N=242,828). For all participants meeting these eligibility criteria, we extracted relevant ICD-10 codes and the corresponding date of documentation for 58 chronic conditions (Table 1) cataloged in the Centers for Medicare & Medicaid Services (CMS) Chronic Conditions Data Warehouse (5). Due to reliance on ICD-10 codes for both cohort identification and as the source of condition information, we limited participant visits to on or after October 1, 2015, i.e., initiation of ICD-10 implementation. Thus, data included in this analysis are from October 1, 2015, to July 1, 2022, i.e., latest date available in the version 7 release.

**Table 1.**
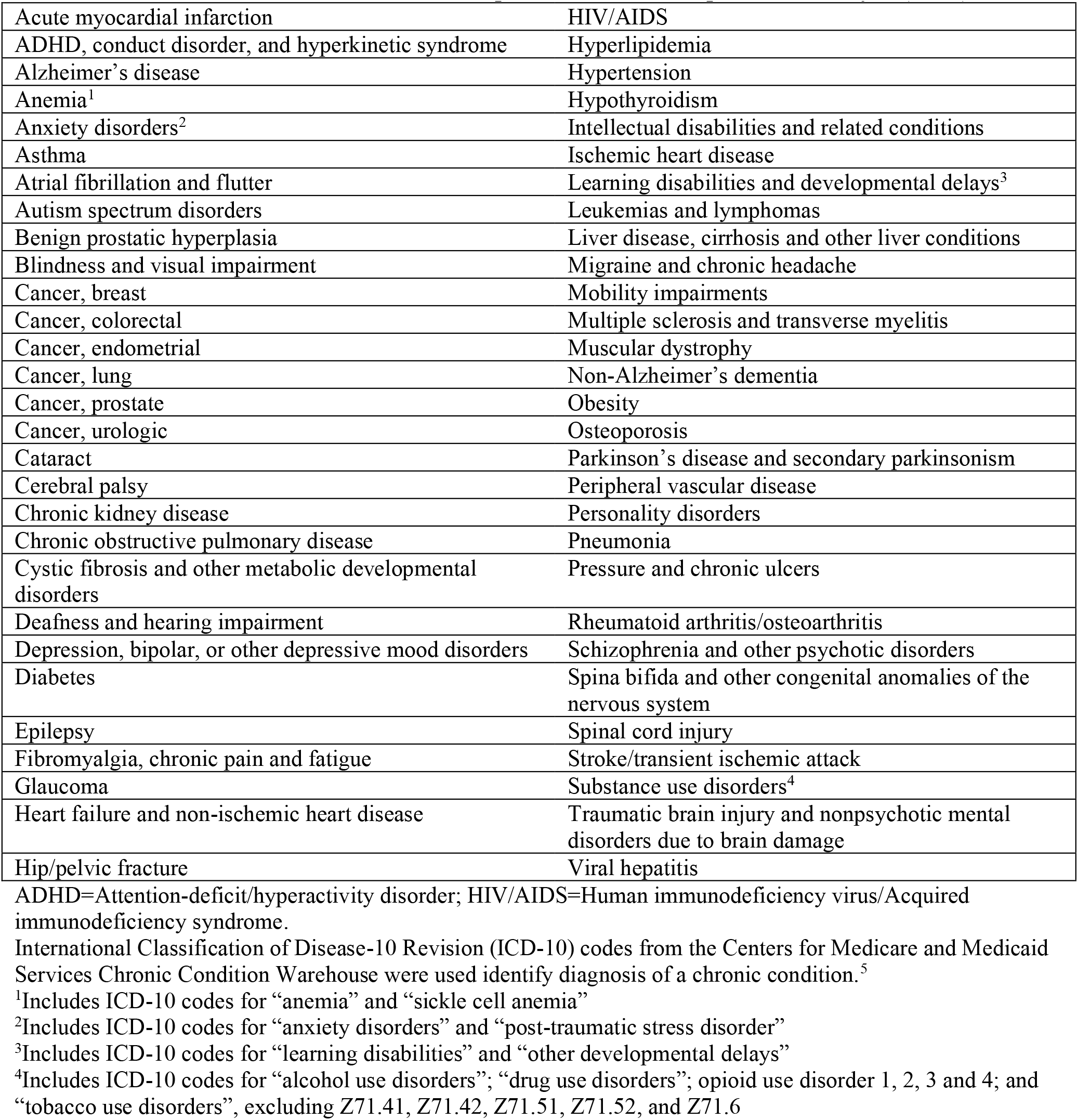
Chronic conditions included in the multiple chronic condition prevalence analysis (N=58)

|  |  |
| --- | --- |
| Acute myocardial infarction | HIV/AIDS |
| ADHD, conduct disorder, and hyperkinetic syndrome | Hyperlipidemia |
| Alzheimer's disease | Hypertension |
| Anemia<sup>1</sup> | Hypothyroidism |
| Anxiety disorders<sup>2</sup> | Intellectual disabilities and related conditions |
| Asthma | Ischemic heart disease |
| Atrial fibrillation and flutter | Learning disabilities and developmental delays<sup>3</sup> |
| Autism spectrum disorders | Leukemias and lymphomas |
| Benign prostatic hyperplasia | Liver disease, cirrhosis and other liver conditions |
| Blindness and visual impairment | Migraine and chronic headache |
| Cancer, breast | Mobility impairments |
| Cancer, colorectal | Multiple sclerosis and transverse myelitis |
| Cancer, endometrial | Muscular dystrophy |
| Cancer, lung | Non-Alzheimer's dementia |
| Cancer, prostate | Obesity |
| Cancer, urologic | Osteoporosis |
| Cataract | Parkinson's disease and secondary parkinsonism |
| Cerebral palsy | Peripheral vascular disease |
| Chronic kidney disease | Personality disorders |
| Chronic obstructive pulmonary disease | Pneumonia |
| Cystic fibrosis and other metabolic developmental disorders | Pressure and chronic ulcers |
| Deafness and hearing impairment | Rheumatoid arthritis/osteoarthritis |
| Depression, bipolar, or other depressive mood disorders | Schizophrenia and other psychotic disorders |
| Diabetes | Spina bifida and other congenital anomalies of the nervous system |
| Epilepsy | Spinal cord injury |
| Fibromyalgia, chronic pain and fatigue | Stroke/transient ischemic attack |
| Glaucoma | Substance use disorders<sup>4</sup> |
| Heart failure and non-ischemic heart disease | Traumatic brain injury and nonpsychotic mental disorders due to brain damage |
| Hip/pelvic fracture | Viral hepatitis |

ADHD=Attention-deficit/hyperactivity disorder; HIV/AIDS=Human immunodeficiency virus/Acquired immunodeficiency syndrome.
International Classification of Disease-10 Revision (ICD-10) codes from the Centers for Medicare and Medicaid Services Chronic Condition Warehouse were used identify diagnosis of a chronic condition.<sup>5</sup>
<sup>1</sup>Includes ICD-10 codes for "anemia" and "sickle cell anemia"
<sup>2</sup>Includes ICD-10 codes for "anxiety disorders" and "post-traumatic stress disorder"
<sup>3</sup>Includes ICD-10 codes for "learning disabilities" and "other developmental delays"
<sup>4</sup>Includes ICD-10 codes for "alcohol use disorders"; "drug use disorders"; opioid use disorder 1, 2, 3 and 4; and "tobacco use disorders", excluding Z71.41, Z71.42, Z71.51, Z71.52, and Z71.6

Then, we used the Python programming language in Jupyter Notebook to chronologically order the chronic condition ICD-10 codes by date of documentation for each participant. By distinguishing the onset date of the second unique chronic condition, we were able to identify participants with MCCs. We calculated the number of participants with 0, 1, 2, 3, 4, 5, and 6+ chronic conditions in the overall cohort and stratified by age category, i.e., early adulthood (18-39 years), middle adulthood (40-49 years), late-middle adulthood (50-64 years), late adulthood (65-74 years), and advanced old age (75-89 years) (6,7). We used the age at first documentation of the second chronic condition for categorization. Finally, we calculated the frequency of unique MCC condition combinations by chronic condition count (i.e., 2, 3, 4, 5, and 6+) and age category.

## Results

Approximately 76% (n=183,753) of participants were diagnosed with MCCs, with over 40% of participants (n=98,885) diagnosed with 6 or more conditions (Table 2). Of the remaining participants, 11% (n=27,122) were diagnosed with a single chronic condition, and 13% (n=31,953) did not have a chronic condition documented. Overall, participants (Table 3) were on average 49.8 (SD=16.7) years of age and primarily women (60.9%), White (54.6%), not Hispanic or Latino (77.3%), married/partnered (49.9%), highly educated (68.7% with some college or higher), insured (92.2%), and not currently employed (47.7%). Compared to participants diagnosed with 0 or 1 conditions, participants diagnosed with MCC were older (mean age of 52.8 years vs. 40.4 years). In addition, a higher percentage of participants with MCCs were men (37.4% vs. 33.8%), White (58.6% vs. 49.6%), not Hispanic/Latino (79.1% vs. 71.4%), insured (93.1% vs. 89.5%), ever married (75.7% vs. 66.4%), and not currently employed (52.3% vs. 33.5%), with a much greater frequency of retired participants (26.4% vs. 10.2%), compared to participants diagnosed with 0 or 1 conditions. The percentage of participants within an age category diagnosed with 6 or more chronic conditions increases with age (Table 2), from 33.78% in early adulthood to 68.04% in advanced old age.

**Table 2.** Number of participants with multiple chronic conditions overall and by age categorization.

| **Condition count** | **n** | **%** |
| --- | --- | --- |
| **Overall (N=183,753)** |  |  |
| 2 | 24,039 | 9.90 |
| 3 | 21,910 | 9.02 |
| 4 | 20,075 | 8.27 |
| 5 | 18,844 | 7.76 |
| 6+ | 98,885 | 40.72 |
| **Early adulthood, 18-39 years (n=40,237)** |  |  |
| 2 | 9,327 | 23.18 |
| 3 | 7235 | 17.98 |
| 4 | 5,648 | 14.04 |
| 5 | 4,435 | 11.02 |
| 6+ | 13,592 | 33.78 |
| **Middle adulthood, 40-49 years (n=28,906)** |  |  |
| 2 | 4,139 | 14.32 |
| 3 | 3,689 | 12.76 |
| 4 | 3,244 | 11.22 |
| 5 | 3,052 | 10.56 |
| 6+ | 14,782 | 51.14 |
| **Late middle adulthood, 50-64 years (n=66,705)** |  |  |
| 2 | 6,786 | 10.17 |
| 3 | 6,945 | 10.41 |
| 4 | 6,895 | 10.34 |
| 5 | 6,715 | 10.07 |
| 6+ | 39,364 | 59.01 |
| **Late adulthood, 65-74 years (n=35,549)** |  |  |
| 2 | 2,778 | 7.81 |
| 3 | 3,019 | 8.49 |
| 4 | 3,227 | 9.08 |
| 5 | 3,462 | 9.74 |
| 6+ | 23,063 | 64.88 |
| **Advanced old age, 75-89 years (n= 11,289)** |  |  |
| 2 | 811 | 7.18 |
| 3 | 844 | 7.48 |
| 4 | 919 | 8.14 |
| 5 | 1,034 | 9.16 |
| 6+ | 7,681 | 68.04 |

**Table 3.**
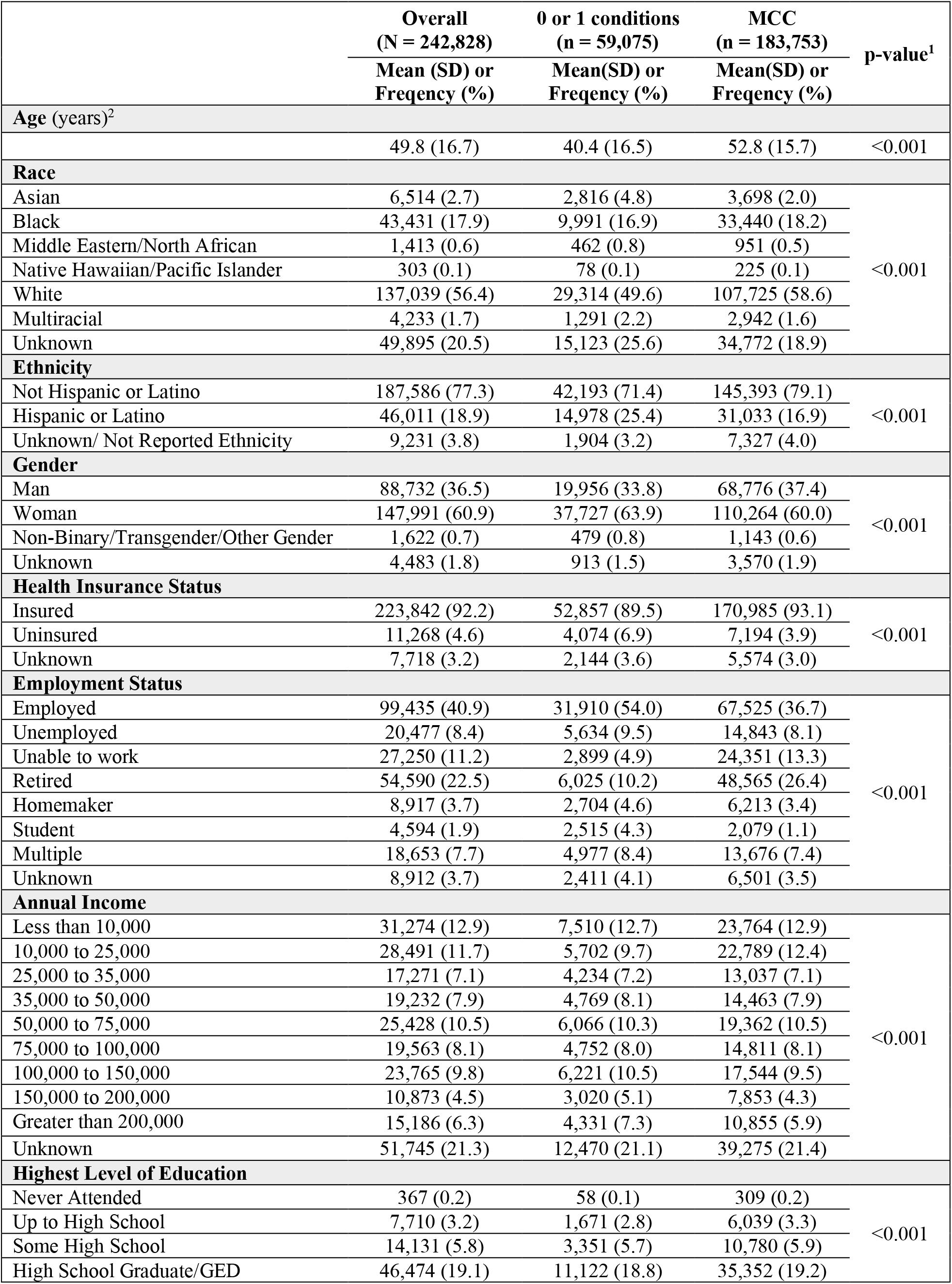
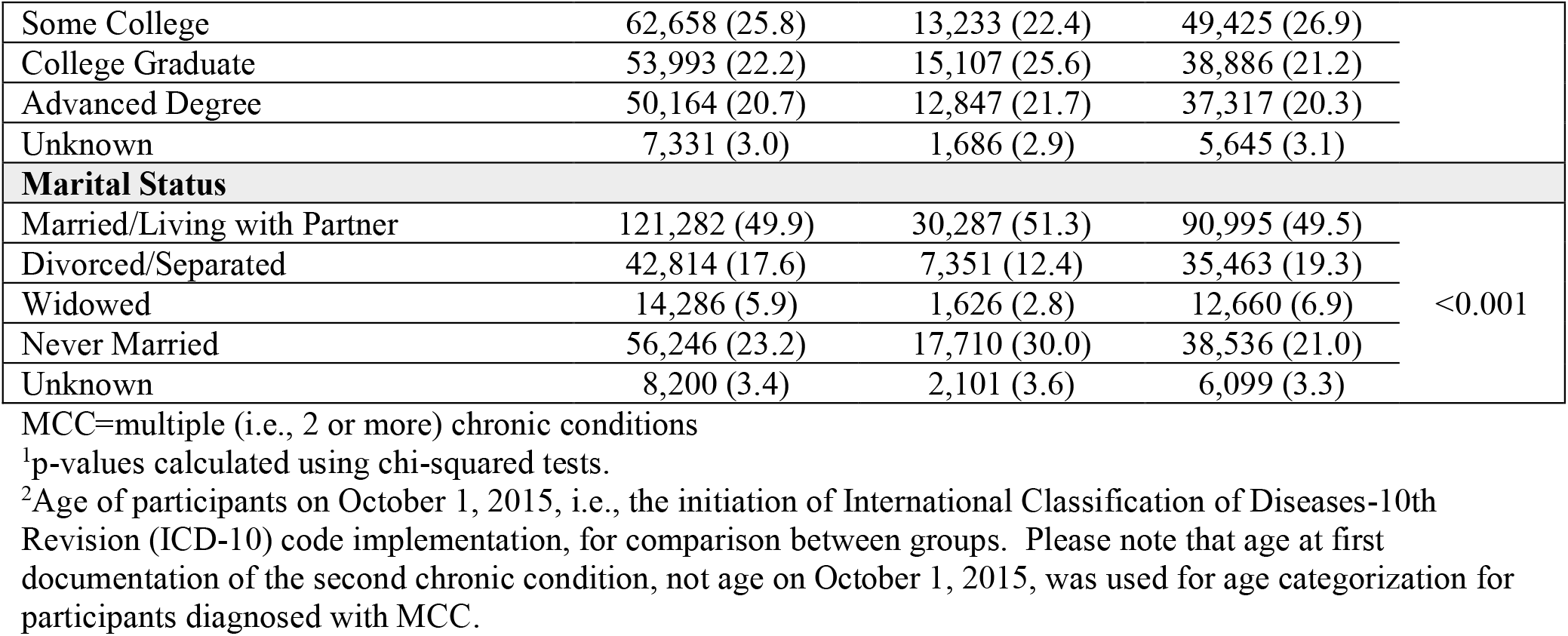
Participant characteristics (N=242,828)

|  | **Overall**<br>**(N = 242,828)** | **0 or 1 conditions**<br>**(n = 59,075)** | **MCC**<br>**(n = 183,753)** | **p-value<sup>1</sup>** |
| --- | --- | --- | --- | --- |
|  | **Mean (SD) or**<br>**Frequency (%)** | **Mean(SD) or**<br>**Frequency (%)** | **Mean(SD) or**<br>**Frequency (%)** |  |
| **Age (years)<sup>2</sup>** |  |  |  |  |
|  | 49.8 (16.7) | 40.4 (16.5) | 52.8 (15.7) | <0.001 |
| **Race** |  |  |  |  |
| Asian | 6,514 (2.7) | 2,816 (4.8) | 3,698 (2.0) | <0.001 |
| Black | 43,431 (17.9) | 9,991 (16.9) | 33,440 (18.2) |  |
| Middle Eastern/North African | 1,413 (0.6) | 462 (0.8) | 951 (0.5) |  |
| Native Hawaiian/Pacific Islander | 303 (0.1) | 78 (0.1) | 225 (0.1) |  |
| White | 137,039 (56.4) | 29,314 (49.6) | 107,725 (58.6) |  |
| Multiracial | 4,233 (1.7) | 1,291 (2.2) | 2,942 (1.6) |  |
| Unknown | 49,895 (20.5) | 15,123 (25.6) | 34,772 (18.9) |  |
| **Ethnicity** |  |  |  |  |
| Not Hispanic or Latino | 187,586 (77.3) | 42,193 (71.4) | 145,393 (79.1) | <0.001 |
| Hispanic or Latino | 46,011 (18.9) | 14,978 (25.4) | 31,033 (16.9) |  |
| Unknown/ Not Reported Ethnicity | 9,231 (3.8) | 1,904 (3.2) | 7,327 (4.0) |  |
| **Gender** |  |  |  |  |
| Man | 88,732 (36.5) | 19,956 (33.8) | 68,776 (37.4) | <0.001 |
| Woman | 147,991 (60.9) | 37,727 (63.9) | 110,264 (60.0) |  |
| Non-Binary/Transgender/Other Gender | 1,622 (0.7) | 479 (0.8) | 1,143 (0.6) |  |
| Unknown | 4,483 (1.8) | 913 (1.5) | 3,570 (1.9) |  |
| **Health Insurance Status** |  |  |  |  |
| Insured | 223,842 (92.2) | 52,857 (89.5) | 170,985 (93.1) | <0.001 |
| Uninsured | 11,268 (4.6) | 4,074 (6.9) | 7,194 (3.9) |  |
| Unknown | 7,718 (3.2) | 2,144 (3.6) | 5,574 (3.0) |  |
| **Employment Status** |  |  |  |  |
| Employed | 99,435 (40.9) | 31,910 (54.0) | 67,525 (36.7) | <0.001 |
| Unemployed | 20,477 (8.4) | 5,634 (9.5) | 14,843 (8.1) |  |
| Unable to work | 27,250 (11.2) | 2,899 (4.9) | 24,351 (13.3) |  |
| Retired | 54,590 (22.5) | 6,025 (10.2) | 48,565 (26.4) |  |
| Homemaker | 8,917 (3.7) | 2,704 (4.6) | 6,213 (3.4) |  |
| Student | 4,594 (1.9) | 2,515 (4.3) | 2,079 (1.1) |  |
| Multiple | 18,653 (7.7) | 4,977 (8.4) | 13,676 (7.4) |  |
| Unknown | 8,912 (3.7) | 2,411 (4.1) | 6,501 (3.5) |  |
| **Annual Income** |  |  |  |  |
| Less than 10,000 | 31,274 (12.9) | 7,510 (12.7) | 23,764 (12.9) | <0.001 |
| 10,000 to 25,000 | 28,491 (11.7) | 5,702 (9.7) | 22,789 (12.4) |  |
| 25,000 to 35,000 | 17,271 (7.1) | 4,234 (7.2) | 13,037 (7.1) |  |
| 35,000 to 50,000 | 19,232 (7.9) | 4,769 (8.1) | 14,463 (7.9) |  |
| 50,000 to 75,000 | 25,428 (10.5) | 6,066 (10.3) | 19,362 (10.5) |  |
| 75,000 to 100,000 | 19,563 (8.1) | 4,752 (8.0) | 14,811 (8.1) |  |
| 100,000 to 150,000 | 23,765 (9.8) | 6,221 (10.5) | 17,544 (9.5) |  |
| 150,000 to 200,000 | 10,873 (4.5) | 3,020 (5.1) | 7,853 (4.3) |  |
| Greater than 200,000 | 15,186 (6.3) | 4,331 (7.3) | 10,855 (5.9) |  |
| Unknown | 51,745 (21.3) | 12,470 (21.1) | 39,275 (21.4) |  |
| **Highest Level of Education** |  |  |  |  |
| Never Attended | 367 (0.2) | 58 (0.1) | 309 (0.2) | <0.001 |
| Up to High School | 7,710 (3.2) | 1,671 (2.8) | 6,039 (3.3) |  |
| Some High School | 14,131 (5.8) | 3,351 (5.7) | 10,780 (5.9) |  |
| High School Graduate/GED | 46,474 (19.1) | 11,122 (18.8) | 35,352 (19.2) |  |
| Some College | 62,658 (25.8) | 13,233 (22.4) | 49,425 (26.9) |  |
| College Graduate | 53,993 (22.2) | 15,107 (25.6) | 38,886 (21.2) |  |
| Advanced Degree | 50,164 (20.7) | 12,847 (21.7) | 37,317 (20.3) |  |
| Unknown | 7,331 (3.0) | 1,686 (2.9) | 5,645 (3.1) |  |
| **Marital Status** |  |  |  |  |
| Married/Living with Partner | 121,282 (49.9) | 30,287 (51.3) | 90,995 (49.5) |  |
| Divorced/Separated | 42,814 (17.6) | 7,351 (12.4) | 35,463 (19.3) |  |
| Widowed | 14,286 (5.9) | 1,626 (2.8) | 12,660 (6.9) | <0.001 |
| Never Married | 56,246 (23.2) | 17,710 (30.0) | 38,536 (21.0) |  |
| Unknown | 8,200 (3.4) | 2,101 (3.6) | 6,099 (3.3) |  |

MCC=multiple (i.e., 2 or more) chronic conditions
<sup>1</sup>p-values calculated using chi-squared tests.
<sup>2</sup>Age of participants on October 1, 2015, i.e., the initiation of International Classification of Diseases-10th Revision (ICD-10) code implementation, for comparison between groups. Please note that age at first documentation of the second chronic condition, not age on October 1, 2015, was used for age categorization for participants diagnosed with MCC.

The top five most frequently diagnosed MCC condition combinations in each age category are detailed in Table 4. The most frequently occurring combination in each age categorization was as follows: early adulthood – anxiety disorders and depression, bipolar, or other depressive mood disorders (n=845); middle and late middle adulthood – fibromyalgia, chronic pain and fatigue and rheumatoid arthritis/osteoarthritis (n=204 and n=457, respectively); and late adulthood and advanced old age – hyperlipidemia and hypertension (n=200 and n=59, respectively).

**Table 4.** Most frequently diagnosed multiple chronic condition combinations in each age category.

| **Condition combinations** | **n** |
| --- | --- |
| **Early adulthood, 18-39 years (n=40,237)** |  |
| Anxiety disorders; Depression, bipolar, or other depressive mood disorders | 845 |
| Anemia; Obesity | 355 |
| Anxiety disorders; Depression, bipolar, or other depressive mood disorders; Substance use disorders | 340 |
| Anxiety disorders; Fibromyalgia, chronic pain and fatigue | 238 |
| Anxiety disorders; Depression, bipolar, or other depressive mood disorders; Fibromyalgia, chronic pain and fatigue | 216 |
| **Middle adulthood, 40-49 years (n=28,906)** |  |
| Fibromyalgia, chronic pain and fatigue; Rheumatoid arthritis/osteoarthritis | 204 |
| Anxiety disorders; Depression, bipolar, or other depressive mood disorders | 168 |
| Hyperlipidemia; Hypertension | 104 |
| Hypertension; Substance use disorders | 103 |
| Hypertension; Obesity | 84 |
| **Late middle adulthood, 50-64 years (n=66,705)** |  |
| Fibromyalgia, chronic pain and fatigue; Rheumatoid arthritis/osteoarthritis | 457 |
| Hyperlipidemia; Hypertension | 385 |
| Hypertension; Substance use disorders | 216 |
| Diabetes; Hyperlipidemia; Hypertension | 204 |
| Diabetes; Hypertension | 173 |
| **Late adulthood, 65-74 years (n=35,549)** |  |
| Hyperlipidemia; Hypertension | 200 |
| Fibromyalgia, chronic pain and fatigue; Rheumatoid arthritis/osteoarthritis | 182 |
| Cataract; Glaucoma | 94 |
| Hyperlipidemia; Hypertension; Rheumatoid arthritis/osteoarthritis | 85 |
| Hypertension; Rheumatoid arthritis/osteoarthritis | 84 |
| **Advanced old age, 75-89 years (n=11,289)** |  |
| Hyperlipidemia; Hypertension | 59 |
| Fibromyalgia, chronic pain and fatigue; Rheumatoid arthritis/osteoarthritis | 46 |
| Cataract; Glaucoma | 44 |
| Hyperlipidemia; Hypertension; Ischemic heart disease | 38 |
| Diabetes; Hyperlipidemia; Hypertension | 26 |

## Discussion

We estimated MCC prevalence in adults living in the U.S. from early adulthood to advanced old age using ICD-10 codes from participants enrolled in *AoU*. We found that approximately 76% of participants have MCC, with higher prevalence as age increases. These findings emphasize the value of *AoU* in identification of MCC burden throughout adulthood. We believe the percentage of adults with MCCs in this study is higher than previous reports due to our robust inclusion of 58 chronic conditions, selected based on the CMS Chronic Conditions Data Warehouse. The characteristics of adult participants who are enrolled in *AoU* and consented to share EHR data – i.e., women, White, not Hispanic/Latino, married/partnered, highly educated, insured, and not currently employed – must also be considered when interpreting the reported estimates and generalizing results to the broader U.S., and worldwide, population.

While sampling bias should be taken into account, this analysis contributes to our understanding of the impact of MCCs. Given the notable prevalence of MCCs across adulthood, there is a continued need for innovative care modalities and public health initiatives to address the multifaceted nature of MCC (8). MCC are associated with clinical complexity characterized by increased symptom burden, high health care utilization and cost, and extensive caregiver strain (9). Ongoing research efforts that seek to both reduce the burden of MCCs on the healthcare system and the development of person- and family-centered care strategies across the healthcare continuum are needed (9).

We found that the most frequent MCC condition combinations vary by age category. Notably, a combination of mental, rather than physical, health conditions (i.e., anxiety disorders and depression, bipolar, or other depressive mood disorders) was the most common MCC combination in early adulthood. Following early adulthood, combinations of physical health conditions were the most common (e.g., hyperlipidemia and hypertension in late adulthood and advanced old age). For precision health, characterizing MCC profiles and the most frequently co-occurring conditions in different age categories can inform targeted prevention and treatment approaches and allow clinicians to better anticipate and manage comorbidities in their patients (10).

In summary, this brief report fills an important knowledge gap and provides a foundation for future studies on MCC correlates, predictors, and outcomes by communicating current MCC prevalence estimates, that capture a wider range of chronic conditions, across the adult lifespan.

## Data Availability

All data are available with registration through https://www.researchallofus.org/.

## Acknowledgements

Research reported in this publication was supported by the National Institute of Nursing Research of the National Institutes of Health under Award Numbers K01NR020504, R00NR017651, and T32008857. The *All of Us* Research Program would not be possible without the partnership of its participants.

## Data availability

No new data were generated or analyzed in support of this brief report. Researchers interested in registering for access to the All of Us Research Program data can visit: https://www.researchallofus.org/

